# Mediating effect of pro-inflammatory cytokines in the association between depression, anxiety, and cardiometabolic disorders in an ethnically diverse community-dwelling middle-aged and older US population

**DOI:** 10.1101/2025.04.14.25325836

**Authors:** Asma Hallab, The Health and Aging Brain Study (HABS-HD) Study Team

**Affiliations:** Neuroimaging Laboratory - Psychiatry and Radiology Departments – Mass General Brigham - Harvard Medical School, Boston, Massachusetts, USA; Biologie Intégrative et Physiologie (BIP) – Parcours Neurosciences Cellulaires et Integrées. Faculté des Sciences et Ingénierie, Sorbonne Université, Paris, France; Charité - Universitätsmedizin Berlin, Corporate member of Freie Universität Berlin and Humboldt-Universität zu Berlin. Berlin, Germany; Pathologies du Sommeil. Faculté de Médecine Pitié-Salpêtrière, Sorbonne Université, Paris, France

**Author notes:** **Corresponding author:** Dr. med. Dr. Asma Hallab. Charité Universitätsmedizin – Berlin. Chariteplatz 1, 10117 Berlin, Germany. **Conflict of interests:** None. **HABS-HD MPIs:** Sid E O’Bryant, Kristine Yaffe, Arthur Toga, Robert Rissman, & Leigh Johnson; and the HABS-HD Investigators: Meredith Braskie, Kevin King, James R Hall, Melissa Petersen, Raymond Palmer, Robert Barber, Yonggang Shi, Fan Zhang, Rajesh Nandy, Roderick McColl, David Mason, Bradley Christian, Nicole Phillips, Stephanie Large, Joe Lee, Badri Vardarajan, Monica Rivera Mindt, Amrita Cheema, Lisa Barnes, Mark Mapstone, Annie Cohen, Amy Kind, Ozioma Okonkwo, Raul Vintimilla, Zhengyang Zhou, Michael Donohue, Rema Raman, Matthew Borzage, Michelle Mielke, Beau Ances, Ganesh Babulal, Jorge Llibre-Guerra, Carl Hill and Rocky Vig.

**Keywords:** **<u>Keywords</u>**: Aging, Neuroinflammation, Cardiovascular risk, Ethnic disparity, Tumor Necrosis Factor-Alpha, Interleukin-6

## Abstract

**Introduction:** Neuroinflammation is associated with depression and anxiety risk, both of which demonstrate a bilateral relationship with cardiometabolic disorders. Systemic inflammation is also commonly described in patients with cardiometabolic disorders. It is, thus, unclear whether pro-inflammatory cytokines might mediate the relationship between depression, anxiety, and cardiometabolic disorders, particularly in advanced ages.

**Methods:** The multiethnic ≥ 50-year-old study population is a subset of the Health and Aging Brain Study: Health Disparities (HABS-HD). Adjusted logistic and linear regression models were applied to assess associations. Non-linearity was evaluated using restricted cubic splines. Statistical mediation analysis was used to determine the role of inflammation (Tumor Necrosis Factor-alpha (TNF-alpha) and Interleukin-6 (IL-6)). Models were corrected for multiple testing using the False Discovery Rate (FDR)-method.

**Results:** In the 2,093 included cases, depression and/or anxiety were significantly associated with 62% higher odds of Cardiovascular Disorder (CVD) (OR=1.62 [95% CI: 1.22-2.15]), 54% of type 2 diabetes (T2DM) (OR=1.54 [95% CI: 1.29-1.85]), 26% of hypertension (OR=26% [95% CI: 1.07-1.48]), and 29% of obesity (OR=1.29 [95% CI: 1.11-1.51]).

Only IL-6 showed a significant mediating role in the association of depression and/or anxiety with CVD (10%, *p-value_FDR_*=0.016), T2DM (13%, *p-value_FDR_*<0.001), hypertension (16%, *p-value_FDR_*<0.001), and obesity (23%, *p-value_FDR_*<0.001).

**Conclusions:** Depression and anxiety were significantly associated with higher odds of major cardiometabolic disorders, and IL-6 partly mediated these associations. Clinical studies are needed to replicate the findings and specifically cluster high-risk profiles.

## 1. Introduction

Depression and anxiety are the most prevalent mental health disorders worldwide and have a high impact on the quality of life of affected persons. (1) They are also associated with higher morbidity and mortality rates across different age groups. (1) Several risk factors are involved in their pathophysiology and evolution. Childhood adversities, (2, 3) chronic emotional stress, (4) traumatic events, (5) and predisposing mental illnesses, be of psychiatric, (6) neurodevelopmental, (7) or neurological (8, 9) nature, might all present favorable grounds for developing a depressive or anxiety disorder.

Recent studies have highlighted the role of neuroinflammation in the genesis of mental disorders through increased activation of microglia and astrocytes and the secretion of pro-inflammatory cytokines. (10) Elevated levels of Tumor Necrosis Factor (TNF)-alpha and Interleukin (IL)-6 are particularly associated with depression and/or anxiety by disrupting the production and function of the neurotransmitters. (11–13) Similarly, systemic low-grade inflammation can induce comparable effects after breaching the blood-brain barrier and majoring the underlying neuroinflammatory process. (14) (Neuro-)Inflammation is also associated with structural (15) and functional (16) remodeling of the brain structures and hyperactivation of the hypothalamic-pituitary-adrenal (HPA) axis, aggravating the systemic stress reaction. (13, 17, 18)

The association between neuroinflammation and depression is bidirectional, complex, and involves intricate mechanisms. (10) While there is stronger evidence of the association between systemic inflammatory biomarkers and depression, (19, 20) fewer studies reported similar observations in patients with anxiety and stress disorders. (21) Noteworthy is, thus, the high comorbidity between depression and anxiety and their mutual interactions. (22)

Epidemiological data are also in favor of a significant association between cardiometabolic disorders, depression, and anxiety. People with cardiovascular diseases (CVD) are at higher risk of developing depression and vice versa. (23) Similarly, there is a significant association between diabetes and depression, as depression predicts the onset of type 2 diabetes mellitus (T2DM) (24), and patients with T2DM tend to develop depression in the course of their disease. (25) Anxiety was also highly reported in patients with hypertension (26) and obesity. (27)

The mechanisms through which mental health disorders might interact with cardiometabolic disorders are still not well understood. Published studies explored several coexisting associations independently and without detailing whether a mediation effect can explain the variations. Furthermore, most published data covered young (28–32) and white populations, (20, 33–35) and there is a serious lack of data on ethnic minorities and persons of advanced ages, both of which are considered high-risk groups for depression and anxiety. (36, 37) The main aim of the current study was to assess whether pro-inflammatory cytokines, biomarkers of systemic inflammation, might have, statistically, a significant mediating effect in the association between depression, anxiety, and cardiometabolic disorders.

## 2. Methods

The study was performed following the Strengthening the Reporting of Observational Studies in Epidemiology (STROBE) guidelines. (38)

### 2.1. Study population

The study population was recruited and followed at the Institute of Translational Research at the University of North Texas Health Science Center (UNTHSC) as a subset of the Health and Aging Brain Study: Health Disparities (HABS-HD) project.

The mono-site HABS-HD is a continuum of the Health & Aging Brain among Latino Elders (HABLE) study, which aimed to improve the understanding of Alzheimer’s Disease-related health disparities in older Hispanic adults in the United States (i.e. Mexican Americans) in contrast with non-Hispanic White Americans, and was initiated in September 2017. Distinct from other large-scale clinic-based cohorts, this community-based study recruited participants in community-based events and through media. Snowball recruitment (participants refer others into the study) was regularly observed. (39) The enrollment of an additional 1,000 Black Americans started in February 2021, with a renaming of the cohort to HABS-HD and the actualization of its aims. All participants underwent regular clinical, neuropsychological, biological, and neuroimaging screenings at 24–30-month intervals (interview, functional exam, cognitive battery, physical exam, informant interview, blood draw, proteomics, exosome assays, magnetic resonance imaging, amyloid and tau positron-emission tomography, and consensus classification). (39) A licensed clinician assigned medical research diagnoses based on medical history, medication, clinical laboratory results, and objective measures. Participants were not eligible if they had Type-1 diabetes, severe physical and mental health conditions, active infections, alcohol/substance use disorders, or dementia other than Alzheimer’s type. (39) The statistical analysis of the current study was performed between December 2024 and February 2025 and used data from the 5^th^ release of HABS-HD.

HABS-HD is funded by grants from the National Institute on Aging (NIA) and, in 2020, UNTHSC provided funding for the addition of 1,000 African Americans to the cohort.

All procedures contributing to this work comply with the ethical standards of the relevant national and institutional committees on human experimentation and with the Helsinki Declaration of 1975, as revised in 2013. Ethical approval was obtained from the local institutional reviewing board (North Texas Regional Institutional Review Board). Participants gave written informed consent. The current research is based on a secondary analysis of De-Identified data.

### 2.2. Depression and anxiety

Participants were reported positive for depression or anxiety if they met the following criteria:

**• Depression:** (1) Self-reported medical history of depression OR (2) Self-reported specific medication OR (3) Geriatric Depression total Score (GDS) ≥ 10 points. (40)
**• Anxiety**: (1) Self-reported medical history of anxiety (across the broad spectrum of anxiety disorders), OR (2) self-reported specific medication.

Anxiety and depression are not mutually exclusive, and because of the high comorbidity between them in the cohort, they were integrated into one variable (independent variable).

Seeing the community-based aspect of the study, diagnoses were mainly based on information gathered during interviews, without referring to medical records. Participants were expected to disclose <u>current relevant diagnoses and medications</u>. However, the lack of medical records limits information availability and specificity on the lifetime evolution of disclosed diagnoses. GDS was, however, performed at baseline by a certified trained neuropsychologist.

### 2.3. Cardiometabolic risk factors

Cardiometabolic disorders are reported as binary variables and defined in the study as follows:

**• Cardiovascular disease (CVD)** is defined as a “positive past medical history of heart attack, heart failure, cardiomyopathy, atrial fibrillation, or heart valve replacement”, *<u>OR</u>* “relevant medication”.
**• Type 2 Diabetes Mellitus (T2DM)** if “HbA1c ≥ 6.5 (%)” *<u>OR</u>* “past medical history of diabetes”, *<u>OR</u>* “relevant medication”. As previously mentioned, participants with type 1 diabetes were not eligible for the study.
**• Dyslipidemia** is defined as “Low-Density Lipoprotein (LDL) ≥ 120”, *<u>OR</u>* “Total Cholesterol ≥ 240”, *<u>OR</u>* “Triglycerides (TG) ≥ 200”, *<u>OR</u>* “Past medical history of high Cholesterol”, OR “relevant medication”.
**• Hypertension** is defined as a “past medical history of hypertension”, *<u>OR</u>* “consistent elevation of blood pressure across both measurements”, *<u>OR</u>* “at least two blood pressure readings of Systolic Blood Pressure (SBP) ≥ 140 mmHg or Diastolic Blood Pressure (DBP) ≥ 90 mmHg”, *<u>OR</u>* “relevant medication”.

Furthermore, **Body Mass index (BMI)** is defined as the result of weight (Kg) / height (m)^2^. Fasting levels of total cholesterol, HDL-cholesterol, LDL-cholesterol, and triglyceride were reported in mg/dL. Fasting **HbA1c** was reported in percentage (%).

### 2.4. Inflammation

Systemic inflammation was evaluated based on serum levels of the two most reliable cytokines, TNF-alpha and IL-6, measured in the fasting blood serum and reported in pg/mL. Details were previously described in the methodological study. (39)

### 2.5. Statistical analyses

The statistical analysis and data visualization were performed using RStudio version 2024.04.1. Continuous variables were reported in medians with interquartile ranges (IQR), and count variables were reported as numbers with percentages (%). TNF-alpha and IL-6 were log-transformed. Wilcoxon rank sum test (Mann-Whitney U), Pearson’s Chi-squared (*X^2^*) test, and *X^2^* Kruskall-Wallis were applied for group comparison, and the corresponding *p*-values were reported.

Associations were explored using multivariable linear regression models for continuous dependent variables and multivariable logistic regression models for binary dependent variables. Models were adjusted for age (years), sex (“female” vs. “male”), self-reported ethnic group (“non-Hispanic white”, “Hispanic”, or “African American”), educational background (years), binary assessment of the current smoking status, and BMI (except for obesity models). Models were tested for non-linearity using restricted cubic splines, and only relevant outcomes were visualized.

Owing to a large loss of follow-up at 24 months and the low number of incident cases of cardiometabolic disorders reported during the follow-up, the current study followed only a cross-sectional design using baseline data.

Assumptions required for causal inference are generally not completely met in cross-sectional studies, and particularly in the current study, the chronological order of events can not be determined. Therefore, a non-causal statistical mediation analysis was performed to understand the relationship between variables and explain statistical variations. Statistical mediation analysis was performed following a 1000-fold non-parametric bootstrapping method, and the age and sex-adjusted Average Causal Mediation Effects (ACME), Average Direct Effects (ADE), and Total Effect, as well as their corresponding 95% confidential interval (95% *CI*), were visualized. The percentage of mediated effect (rounded value) was calculated as the proportion of ACME of the Total Effect and reported with the corresponding *p*-value. To reduce type I errors in the different outcomes of interest, adjusting for multiple testing was performed using the False Discovery Rate (FDR) method, and resulting *p_FDR_*-values were reported. The significance level of the two-sided *p*- and *p_FDR_*-values was set at 0.05.

## 3. Results

### 3.1. Study population

The study included 2,093 participants aged between 50 and 92 years with a median age of 66 (IQR: 59-72). Among these, 44% disclosed themselves as “White”, 44% as “Hispanic”, and 12% as “Black”. Women represented 62% of the study population. At baseline, 712 (34%) were diagnosed with depression and 360 (17.2%) with anxiety (**Table 1**). Depression and anxiety were simultaneously diagnosed in 281 cases (13.4%), while 431 (20.6%) had depression without anxiety, and 79 (3.8%) had only anxiety without depression.

Cases with depression were significantly younger (65 vs. 66 years, *p-value=*0.033), predominantly females (71% vs. 57%, *p-value<*0.001), less educated (13 vs. 14 years, *p-value<*0.001), and had higher BMI (30 vs. 29, *p-value<*0.001). More obesity (53% vs. 43%, *p-value<*0.001), hypertension (67% vs. 62%, *p-value=*0.042), and T2DM (31% vs. 22%, *p-value*<0.001) were reported in cases with depression compared to those without depression. Biologically, only serum Triglyceride (119 vs. 108 mg/dL, *p-value=*0.001) and log IL-6 levels (−0.10 vs. 0.01, *p-value<*0.001) were significantly higher in cases with depression (**Table 1**).

Similarly, cases with anxiety tend to be significantly younger (64 vs. 66 years, *p-value*<0.001), predominantly females (75% vs. 59%, *p-value*<0.001), have higher BMI (30 vs. 29, p-value=0.011) and obesity (52% vs. 45%, *p-value=*0.012). Biologically, serum Triglyceride and log IL-6 levels were also higher in cases with anxiety than those without (119 vs. 110, *p-value=*0.020, and 0.05 vs. −0.07, *p-value*<0.001, respectively). Cases with anxiety tend to be current smokers (8.5% vs. 5.3%, *p-value=*0.015) (**Table 1**). Hispanic and Black participants had significantly higher GDS scores than the white non-Hispanic ones (*p-value _FDR_* <0.001, respectively) (**Figure 1.a.**).

**Figure 1:**
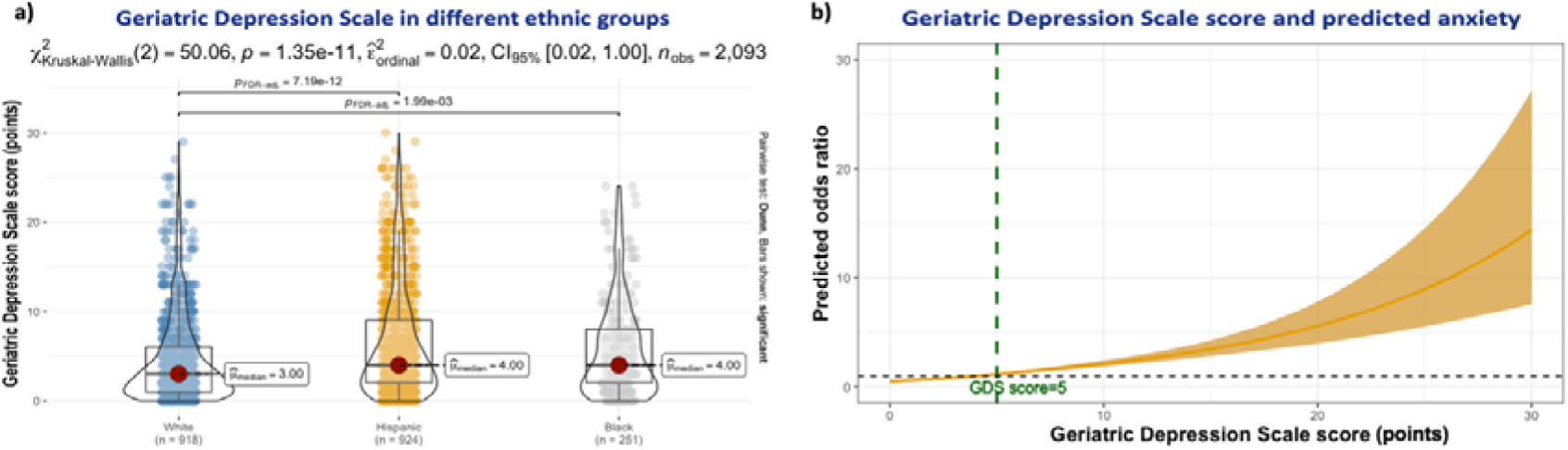
Geriatric Depression Scale and Associations. **1.a.)** Geriatric Depression Scale scores in different ethnic groups: *X^2^* Kruskal-Wallis analysis. **1.b.)** Geriatric Depression Scale score and association with anxiety: Non-linear model.

GDS showed a significant non-linear relationship with the predicted odds of anxiety (*p-value _non-linear_* <0.001), and particularly GDS values higher than the median (five points) were exponentially associated with increased odds of reporting anxiety (**Figure 1.b.**).

TNF-alpha levels showed no significant difference between those with depression and/or anxiety and healthy controls (**Supplementary** Figure 1**.a.**). Participants with depression and/or anxiety had significantly higher BMI (**Supplementary** Figure 1**.b.**), IL-6 (**Supplementary** Figure 1**.c.**), and HbA1c levels (**Supplementary** Figure 1**.d.**).

An increase of one point in GDS score was significantly associated with an increase in HbA1c by 0.03 % (*Adj. ß*= 0.03 (0.02-0.04), *p<0.001*) (**Supplementary** Figure 2**.a.**). An increase of HbA1c by 1% was significantly associated with an increase of GDS by 0.51 points (*Adj. ß*= 0.51 (0.32-0.69), *p<0.001*) (**Supplementary** Figure 2**.b.**).

There was also a significant bilateral association between GDS and BMI, thus with lower coefficients (**Supplementary** Figure 2**.c.** and **Supplementary** Figure 2**.d.**).

### 3.2. Systemic inflammation

Neither TNF-alpha levels were significantly associated with variations in GDS scores, nor GDS scores with variations in TNF-alpha levels (**Figure 1.c. and Figure 1.d.**). In contrast, an increase in log IL-6 was associated with higher GDS scores (*Adj. ß*= 0.71 (0.35-1.1), *p<0.001*) (**Figure 1.e.**). Weaker is the significant association of the GDS score with an increase in log IL-6 (*Adj. ß*= 0.01 (0.00-0.02), *p<0.001*) (**Figure 1.f.**).

**Figure 1:**
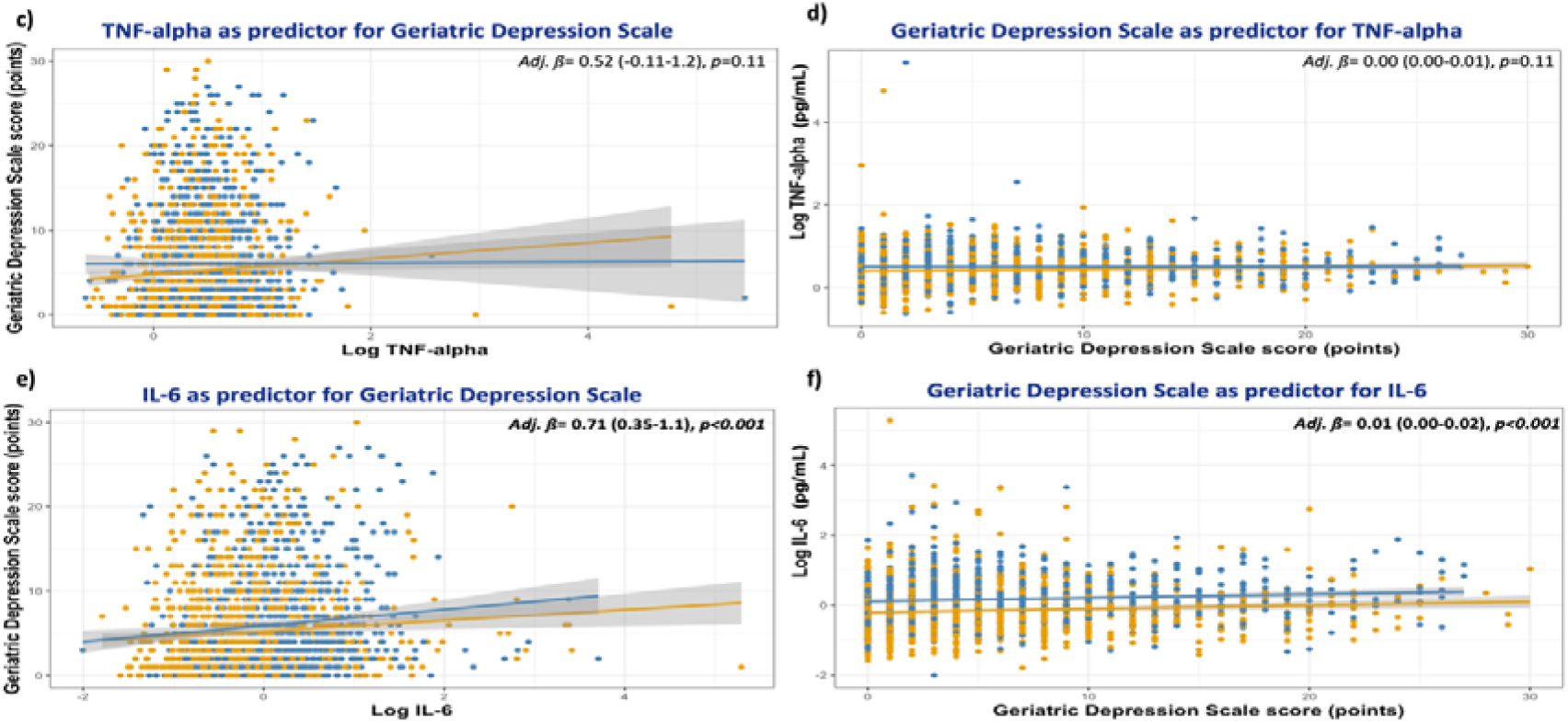
Geriatric Depression Scale and Associations. **1.c.) & 1.d.)** Geriatric Depression Scale score and association with Tumor Necrosis Factor-alpha levels. **1.e.) & 1.f.)** Geriatric Depression Scale score and association with Interleukin-6 levels. <u>Caption</u>: IL-6: Interleukin-6, TNF-alpha: Tumor Necrosis Factor-alpha. Blue: Obese cases with BMI ≥ 30. Yellow: Non-obese cases with BMI < 30.

An increase in Log TNF-alpha was significantly associated with an increase in BMI (*Adj. ß*= 2.9 (2.3-3.6), *p<0.001*), particularly in obese cases (blue) (**Supplementary** Figure 3**.a.**), while an increase in BMI was slightly associated with higher Log TNF-alpha levels (*Adj. ß*= 0.01 (0.01-0.01), *p<0.001*) (**Supplementary** Figure 3**.b.**). Similarly, an increase in log IL-6 levels was significantly associated with an increase in BMI (*Adj. ß*= 3.4 (3.1-3.8), *p<0.001*), particularly in obese cases (blue) (**Supplementary** Figure 3**.c.**). Higher BMI was also significantly associated with a slighter increase in log IL-6 (*Adj. ß*= 0.04 (0.04-0.04), *p<0.001*) (**Supplementary** Figure 3**.d.**).

### 3.3. Depression, anxiety, and cardiometabolic risks

Depression and/or anxiety were significantly associated with 56% higher odds of CVD (OR=1.56 [95% CI: 1.11-2.19]), 49% of T2DM (OR=1.49 [95% CI: 1.19-1.85]), 25% of hypertension (OR=25% [95% CI: 1.02-1.53]), and 37% of obesity (OR=1.37 [95% CI: 1.14-1.65]). The results are visualized in **Figure 2.a.** and summarized in **supplementary table 1**.

**Figure 2:**
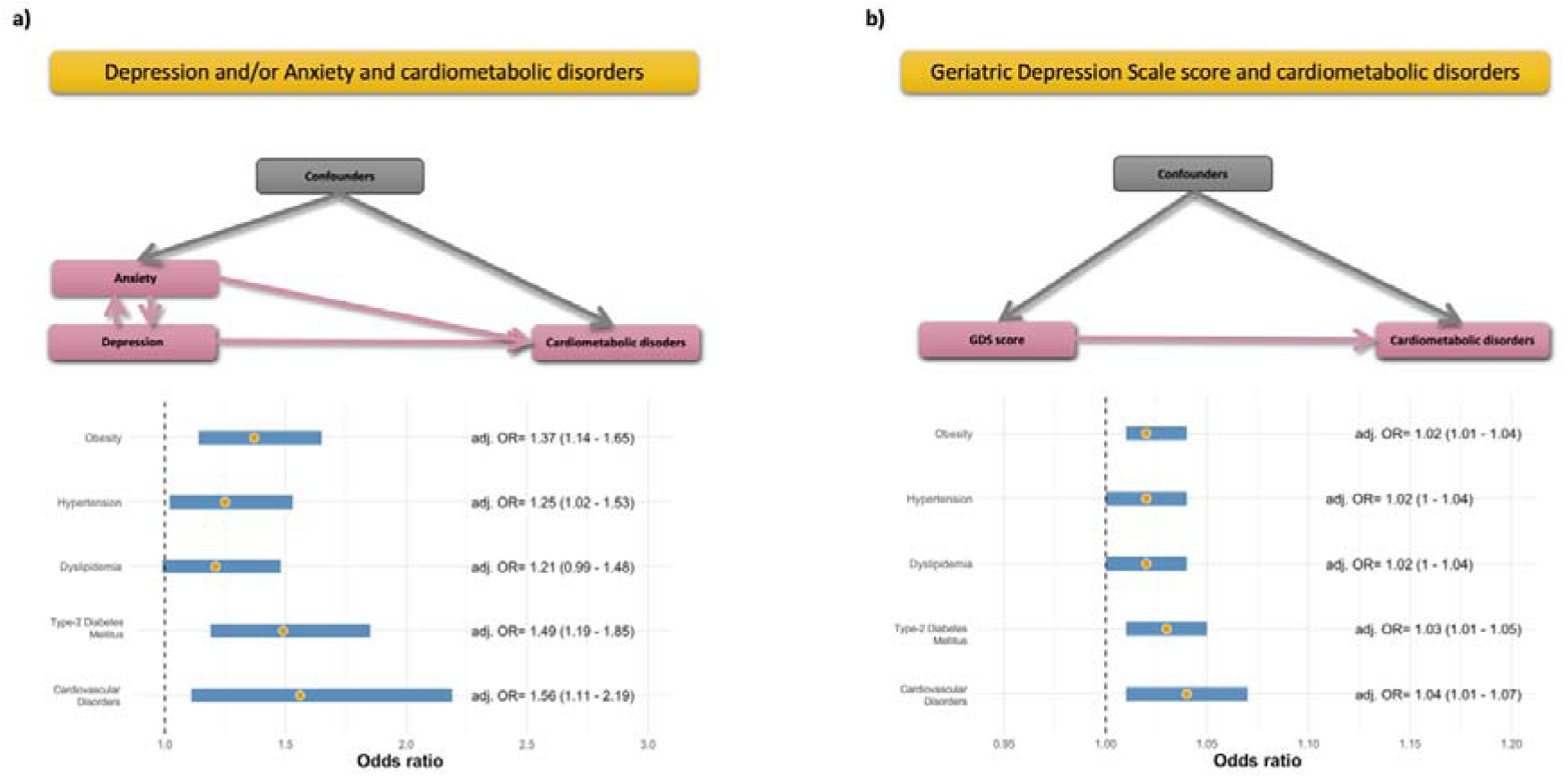
Depression, anxiety, and odds ratios of cardiometabolic disorders. **2.a.)** Depression and/or anxiety, and odds ratios of cardiometabolic disorders. **2.b.)** Geriatric Depression Scale and odds ratios of cardiometabolic disorders.

A sensitivity analysis using GDS as independent continuous variable showed that an increase of one point in GDS score was significantly associated with 4% higher odds of CVD (OR=1.04 [95% CI: 1.01-1.07]), 3% of T2DM (OR=1.03 [95% CI: 1.01-1.05]), 2% of dyslipidemia (OR=1.02 [95% CI: 1.00-1.04]), hypertension (OR=1.02 [95% CI: 1.00-1.04]), and obesity (OR=1.02 [95% CI: 1.01-1.04]). The results are visualized in **Figure 2.b.** and summarized in **supplementary table 2**.

A further sensitivity analysis of results obtained with depression and anxiety, separately, as independent variables can be found in **Supplementary** Figure 4 and detailed in **supplementary Tables 3 and 4**. Depression and anxiety are not mutually exclusive. Therefore, the anxiety analysis included cases with comorbid depression, and the depression analysis included cases with comorbid anxiety. Exploring them separately showed a significant decrease in the statistical power.

### 3.4. Depression, anxiety, and the mediating effect of inflammation

TNF-alpha did not show any significant mediating effect in the relationship between depression and/or anxiety and different cardiometabolic disorders (**Figure 3.a.**).

**Figure 3:**
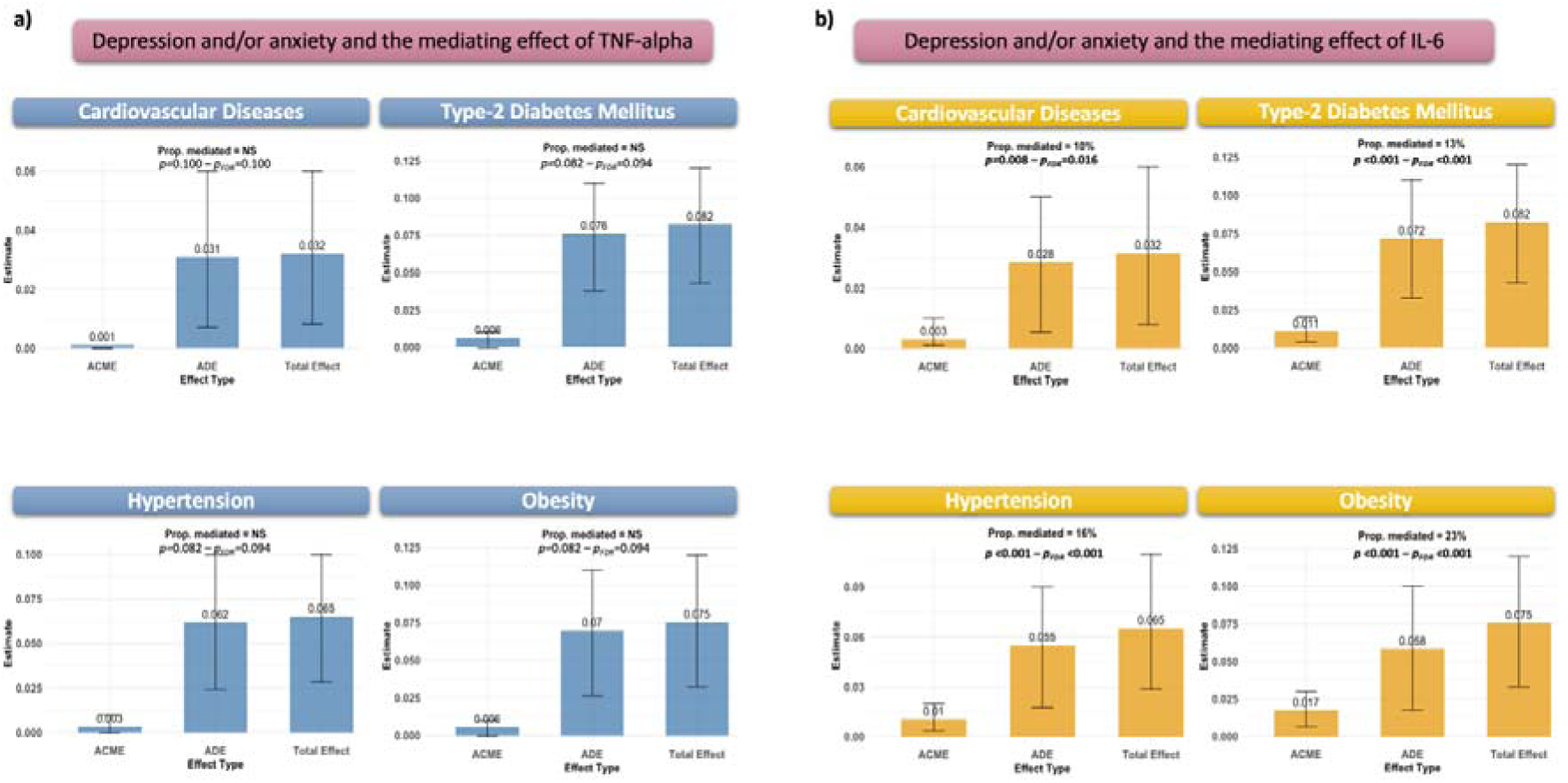
Visualization of mediating analysis. **3.a.)** Tumor Necrosis Factor-alpha as mediator. **3.b.)** Interleukin-6 as mediator.

Il-6 statistically mediated 10% (*p-value _FDR_*=0.016), 13% (*p-value _FDR_*<0.001), 16% (*p-value _FDR_*<0.001), and 23% (*p-value _FDR_*<0.001) of the association between having a depression and/or anxiety and CVD, type 2 diabetes, hypertension, and obesity, respectively. The different associations are visualized in **Figure 3.b.**

## 4. Discussion

The study explored the statistical mediating effect of IL-6 and TNF-alpha in the association between depression and/or anxiety and cardiometabolic disorders in middle-aged and older adults with a multiethnic background. Hispanic and Black participants had significantly higher GDS total scores than non-Hispanic White participants. While the study highlighted a significant association between depression and/or anxiety and higher odds of CVD, T2DM, hypertension, and obesity, IL-6 had a statistical mediating effect ranging between 10% and 23% on these associations. TNF-alpha, however, had a non-significant mediating effect in the association between depression and/or anxiety and the studied cardiometabolic disorders.

- **Depression**

Longitudinal studies are in favor of a bidirectional association between inflammation and depression. While high IL-6 levels predict depression onset after seven years of follow-up, a baseline depressive status predicts, after the same period, consequent CRP elevation. (41) This association starts at an early age and has a long-lasting effect. In children, higher serum IL-6 and CRP levels were associated with higher depression risk in their young adult life. (31) In older adults, higher systemic IL-6 and TNF-alpha levels are associated with suicide risk, (42) reflecting the severity of the underlying depressive symptoms. The association between proinflammatory cytokines and depression and/or anxiety is complex and might be subject to sex-specific patterns. (43) Il-6 is a precursor of CRP (10) and therefore only IL-6 was considered in the current analysis dedicated to pro-inflammatory cytokines IL-6 and TNF-alpha.

- **BMI and obesity**

Depressed patients with higher BMI tend to express higher systemic inflammatory biomarkers. (44) Furthermore, high levels of inflammatory biomarkers and depression are independently associated with less weight loss in patients following bariatric surgery. (45) In pregnant women, preexisting obesity is associated with higher pro-inflammatory cytokine levels and predicts a higher risk of antenatal depression. (46) Similarly, higher levels of proinflammatory cytokine in obese patients predict higher depressive symptoms and lower self-esteem. (44) Overweight and obese antidepressant-naïve adolescents diagnosed with depression have higher severity symptoms, including suicidality, in addition to higher systemic IL-6 compared to the lean control group, (29) and adolescents with MDD have higher TNF-alpha levels than healthy controls. (47) IL-6 is also associated with cognitive dysfunction in adolescents with severe depression and high BMI. (30) Thus, only a few studies explored the simultaneous interaction between depression, obesity, and cytokines. While IL-6 is a significant element in the depression-obesity relationship, (48) the role of TNF-alpha is controversial and still needs to be further explored. (49) The current study failed to show a statistical mediating effect of TNF-alpha on the association with obesity.

- **Diabetes**

The mediating effect of pro-inflammatory cytokines on depression has been demonstrated in patients with type 1 diabetes. (50) However, participants with type 1 diabetes were not included in the source cohort. A few studies explored this association in persons with T2DM and showed inconsistent results. (51) Some genetic variabilities coding for the innate immune system are shared between depression and T2DM. (52) In a biopsychological study, a stress-induced increase in IL-6 levels in patients with T2DM predicted higher depression scores. (53) Furthermore, an improvement in depressive symptoms in patients with T2DM was associated with a decrease in inflammation biomarkers during the follow-up. (54) The current analysis showed significant positive bilateral associations between GDS and HbA1c. Furthermore, IL-6 had a significant statistical mediating effect in the association between depression, anxiety, and T2DM.

- **Dyslipidemia**

Very limited data is available on the association between depression and/or anxiety, neuroinflammation, and dyslipidemia. The current study failed to show robust associations between depression and/or anxiety, and dyslipidemia. Only GDS was a significant predictor of dyslipidemia, but not the primary analysis. Rodent studies have shown that a high-fat-diet induced depression-like behavior in mice and up-regulated Glycerol-3-phosphate acyltransferases 4 (GPAT4)-expression in their Hippocampus. (55) The GPAT4 is expressed in the brain and is responsible for Triacylglycerol synthesis and obesity. In addition to higher lipid levels in mice who received a high-fat diet, their IL-6 and TNF-a levels were significantly higher than the control group. (55) In another study, Simvastatin-based lipid-lowering therapy reduced the inflammatory remodeling in the hippocampus of mice with high-fat-diet-induced depression-like symptoms. (56) Further interventional studies are needed to better understand the particularities of the depression/anxiety-inflammation-dyslipidemia association.

- **Hypertension**

Similarly, there are limited studies on the effect of inflammation on the association between depression and/or anxiety and hypertension. The expression of the norepinephrine transporter (NET) gene might impact IL-6 and TNF-alpha levels and interact with depression and hypertension risks. (57) Furthermore, high IL-6 levels and hypertension were predictors of depression in hemodialyzed patients, while only hypertension predicted anxiety in this same group. (58)

Endothelial dysfunction is associated with hypertension and depression in older adults. (59) In rodent models, the stress-induced depressive-like behavior was associated with an increase in IL-6 and TNF-alpha levels and endothelial dysfunction. (60) Only IL-6 showed a significant mediating relationship in the current study, and no data on potential concomitant endothelial dysfunction are available.

- **Cardiovascular diseases**

The association between depression and CVD is bidirectional, and various underlying mechanisms are implicated. (23) Inflammation might play a pivotal role in bridging the path between both diseases. Large Mendelian randomization studies have demonstrated a causal effect of IL-6 and IL-6 receptors on CVD risk; as a genetic polymorphism responsible for lower IL-6 levels is associated with lower risks of CVD (61, 62) and depression. (19)

Psychotherapeutic treatment for major depressive disorder was significantly associated with improved heart rate variability but not with inflammatory biomarker levels (IL-6, TNF-alpha, and CRP). (63) However, combining physical activity with psychotherapy in the therapeutic management of depression had a beneficial impact on their immunological biomarkers. (64) In the current study, depression and/or anxiety were strongly associated with CVD, and this association was partly mediated by IL-6. It is, therefore, important to acknowledge the importance of mental health in the therapeutic management of cardiovascular diseases and vice versa.

- **Anxiety**

Studies on the association between anxiety, cardiometabolic disorders, and inflammation are limited, non-conclusive, and most of them explored anxiety and depression as a comorbid condition. A large US study on adults aged between 30 and 54 years showed that anxiety, measured using the Trait Anxiety scale of the State-Trait Anxiety Inventory (STAI-T), was neither significantly associated with IL-6, nor CRP levels, in contrast with the significant results using depression scores. Higher STAI-T scores were, thus, significantly associated with higher Triglyceride levels. (65) Another large population study on Swiss adults aged between 35 and 66 found no significant association between anxiety on one side and IL-1ß, IL-6, TNF-alpha, and CRP on the other side. (66) Similarly, a further large population study on Dutch adults with a mean age of 41.9 years showed a significant association between the anxious distress symptom dimension and inflammation index (IL-6 and CRP-based) at baseline but not during the follow-up or in those with concomitant depression. (67) There was also no significant association with the metabolic syndrome index in this same study. (67) A randomized controlled trial showed, however, that inhibiting TNF-alpha activity by administrating Infliximab to patients with baseline CRP levels > 5 mg/L induced a reduction in anxiety symptoms compared to those who received a placebo. (68) Larger clinical studies in anxiety patients, with and without depression, are needed in order to better understand the associations and patterns.

- **Strengths**

The main novelty of the study resides in the large number of included participants of advanced ages and multiethnic backgrounds. This allows for better generalizability of the results and presents a newer perspective contrasting with most published data, mainly based on findings in white and younger participants. The large number of participants with complete data on cardiovascular, metabolic, depressive, and anxiety disorders, and the availability of immunological assessments is a further strength. The mono-cite recruitment of participants allowed for homogeneous assessment strategies and presented an advantage for the data quality. Furthermore, this is the first study to explore the statistical mediating effect of pro-inflammatory cytokines on associations between depression, anxiety, and different cardiometabolic disorders.

- **Limitations**

Despite the novelty of the findings, the study has some limitations. First, the definition of depression and anxiety is mainly based on self-disclosed medical history and medications, and only one test was performed to screen for depression. This might have induced some form of recall bias and does not allow for a solid objectivation of anxiety disorders. GDS total score was, thus, significantly predictive of concomitant anxiety, highlighting the validity of the disclosed diagnoses. While participants might also have disclosed the duration of their mental health disorder, this exact onset date remains uncertain and prone to recall bias, as well. Therefore, the duration of depression and/or anxiety remained outside the scope of the current analysis. Seeing the community-based aspect of the source cohort, diagnoses were mainly based on information gathered during interviews, and no previous medical records were available. Anxiety was broadly assessed without details about sub-classifications. This is a recognized limitation of population- and register-based studies. Clinical studies are, therefore, needed to replicate the findings.

It is also important to recognize that many factors (microbiome of the gastrointestinal tract, stress, vitamin D levels, physical activity, diet, anticoagulants) might influence cytokine levels, leading to between and within individual variabilities. (10, 69) These factors were not controlled for in the analysis, either owing to their non-availability in the source cohort or to simplify the interpretability of the results and reduce the risks of overfitting. There is also no data on the IL-6 and TNF-alpha levels in the CFS of included patients, and the analysis was only based on cytokines as biomarkers of systemic inflammation. Furthermore, the cross-sectional design does not allow for a conclusion on a causal effect, and larger longitudinal population-based studies are needed.

## 5. Conclusions

Depression and/or anxiety are associated with higher odds of cardiometabolic disorders in multiethnic middle-aged and older adults, and systemic inflammation, especially IL-6, plays a significant mediating role in this association. These results highlight the need for more multidisciplinary approaches to manage physical and psychological well-being, particularly in older populations. Clinical studies are needed to replicate the findings and specifically cluster at-high-risk profiles. Further research also needs to explore targeted interventions to reduce neuroinflammation and the consequent impact on cardiovascular risks in patients with depressive and anxiety disorders.

## Declarations

### Ethical approval

All procedures contributing to this work comply with the ethical standards of the relevant national and institutional committees on human experimentation and with the Helsinki Declaration of 1975, as revised in 2013. Ethical approval was obtained from the local institutional reviewing board (North Texas). Participants gave written informed consent.

### Authorization for publication

The principal investigator of the HABS-HD study authorized the publication of the current study.

### Authorship

AH has full access to all of the data and takes responsibility for the integrity of the data and the accuracy of the analysis, visualization, drafting, and editing of the manuscript.

### Data availability

Data can be acquired by qualified researchers after an official request.

## Data Availability

All data produced in the present study are available upon reasonable request to the authors

## Acknowledgment

“Research reported on this publication was supported by the National Institute on Aging of the National Institutes of Health under Award Numbers R01AG054073, R01AG058533, R01AG070862, P41EB015922 and U19AG078109. The content is solely the responsibility of the authors and does not necessarily represent the official views of the National Institutes of Health.”

## Supplementary Figures

**Supplementary Figure 1: Comparison of biological biomarker levels between participants with depression and/or anxiety. 1.a)** Log TNF-alpha (pg/mL). **1.b)** Log IL-6 (pg/mL). 1.c) BMI. 1.d) HbA1c (%).

<u>Caption</u>: BMI: Body Mass Index, HbA1c: Glycated Hemoglobin A1c, IL-6: Interleukin-6, TNF-alpha: Tumor Necrosis Factor-alpha.

**Supplementary Figure 2: Association between geriatric depression scale, BMI, and HbA1c. 2.a.** Geriatric depression scale as predictor for HbA1c. **2.b.** HbA1c as predictor for geriatric depression scale. **2.c.** Geriatric depression scale as predictor for BMI. **2.d.** BMI as predictor for geriatric depression scale.

<u>Caption</u>: BMI: Body Mass Index, HbA1c: Glycated Hemoglobin A1c. Blue: Obese cases with BMI ≥ 30. Yellow: Non-obese cases with BMI < 30.

**Supplementary Figure 3: Association between TNF-alpha, IL-6, and BMI. 3.a.** TNF-alpha as predictor for BMI. **3.b.** BMI as predictor for TNF-alpha. **3.c.** IL-6 as predictor for BMI. **3.d.** as BMI predictor for IL-6.

<u>Caption</u>: BMI: Body Mass Index, IL-6: Interleukin-6, TNF-alpha: Tumor Necrosis Factor-alpha. Blue: Obese cases with BMI ≥ 30. Yellow: Non-obese cases with BMI < 30.

**Supplementary Figure 4: Depression, anxiety, and odds ratios of cardiometabolic disorders. 2.a.)** Depression and odds ratios of cardiometabolic disorders. **2.b.)** Anxiety and odds ratios of cardiometabolic disorders.

<u>Caption</u>: Analysis of depression included also cases of depression with comorbid anxiety, and analysis of anxiety included also cases of anxiety with comorbid depression.

## References

1. Global, regional, and national burden of 12 mental disorders in 204 countries and territories, 1990-2019: a systematic analysis for the Global Burden of Disease Study 2019. Lancet Psychiatry. 2022;9(2):137–50.

2. Hailes HP, Yu R, Danese A, Fazel S. Long-term outcomes of childhood sexual abuse: an umbrella review. Lancet Psychiatry. 2019;6(10):830–9.

3. Macpherson JM, Gray SR, Ip P, McCallum M, Hanlon P, Welsh P, et al. Child maltreatment and incident mental disorders in middle and older ages: a retrospective UK Biobank cohort study. Lancet Reg Health Eur. 2021;11:100224.

4. Racine N, Hetherington E, McArthur BA, McDonald S, Edwards S, Tough S, et al. Maternal depressive and anxiety symptoms before and during the COVID-19 pandemic in Canada: a longitudinal analysis. Lancet Psychiatry. 2021;8(5):405–15.

5. Mesa-Vieira C, Haas AD, Buitrago-Garcia D, Roa-Diaz ZM, Minder B, Gamba M, et al. Mental health of migrants with pre-migration exposure to armed conflict: a systematic review and meta-analysis. Lancet Public Health. 2022;7(5):e469–e81.

6. Bohus M, Stoffers-Winterling J, Sharp C, Krause-Utz A, Schmahl C, Lieb K. Borderline personality disorder. Lancet. 2021;398(10310):1528–40.

7. Lai MC, Kassee C, Besney R, Bonato S, Hull L, Mandy W, et al. Prevalence of co-occurring mental health diagnoses in the autism population: a systematic review and meta-analysis. Lancet Psychiatry. 2019;6(10):819–29.

8. Kwon CS, Rafati A, Ottman R, Christensen J, Kanner AM, Jetté N, et al. Psychiatric Comorbidities in Persons With Epilepsy Compared With Persons Without Epilepsy: A Systematic Review and Meta-Analysis. JAMA Neurol. 2025;82(1):72–84.

9. Liu L, Marshall IJ, Pei R, Bhalla A, Wolfe CD, O’Connell MD, et al. Natural history of depression up to 18 years after stroke: a population-based South London Stroke Register study. Lancet Reg Health Eur. 2024;40:100882.

10. Kiecolt-Glaser JK, Derry HM, Fagundes CP. Inflammation: depression fans the flames and feasts on the heat. Am J Psychiatry. 2015;172(11):1075–91.

11. Roohi E, Jaafari N, Hashemian F. On inflammatory hypothesis of depression: what is the role of IL-6 in the middle of the chaos? J Neuroinflammation. 2021;18(1):45.

12. Liu Y, Ho RC-M, Mak A. Interleukin (IL)-6, tumour necrosis factor alpha (TNF-α) and soluble interleukin-2 receptors (sIL-2R) are elevated in patients with major depressive disorder: A meta-analysis and meta-regression. Journal of Affective Disorders. 2012;139(3):230–9.

13. Chang J, Jiang T, Shan X, Zhang M, Li Y, Qi X, et al. Pro-inflammatory cytokines in stress-induced depression: Novel insights into mechanisms and promising therapeutic strategies. Progress in Neuro-Psychopharmacology and Biological Psychiatry. 2024;131:110931.

14. Varatharaj A, Galea I. The blood-brain barrier in systemic inflammation. Brain, Behavior, and Immunity. 2017;60:1–12.

15. Kang Y, Shin D, Kim A, You SH, Kim B, Han KM, et al. The effect of inflammation markers on cortical thinning in major depressive disorder: A possible mediator of depression and cortical changes. J Affect Disord. 2024;348:229–37.

16. Schmitz CN, Sammer G, Neumann E, Blecker C, Gründer G, Adolphi H, et al. Functional resting state connectivity is differentially associated with IL-6 and TNF-α in depression and in healthy controls. Sci Rep. 2025;15(1):1769.

17. Nikkheslat N, Zunszain PA, Horowitz MA, Barbosa IG, Parker JA, Myint AM, et al. Insufficient glucocorticoid signaling and elevated inflammation in coronary heart disease patients with comorbid depression. Brain Behav Immun. 2015;48:8–18.

18. Lasselin J, Benson S, Hebebrand J, Boy K, Weskamp V, Handke A, et al. Immunological and behavioral responses to in vivo lipopolysaccharide administration in young and healthy obese and normal-weight humans. Brain Behav Immun. 2020;88:283–93.

19. Khandaker GM, Zammit S, Burgess S, Lewis G, Jones PB. Association between a functional interleukin 6 receptor genetic variant and risk of depression and psychosis in a population-based birth cohort. Brain, Behavior, and Immunity. 2018;69:264–72.

20. van Dooren FE, Schram MT, Schalkwijk CG, Stehouwer CD, Henry RM, Dagnelie PC, et al. Associations of low grade inflammation and endothelial dysfunction with depression - The Maastricht Study. Brain Behav Immun. 2016;56:390–6.

21. Buto PT, Shah A, Pearce BD, Lima BB, Almuwaqqat Z, Martini A, et al. Association of systemic inflammation with posttraumatic stress disorder after a myocardial infarction. Brain Behav Immun Health. 2023;30:100629.

22. ter Meulen WG, Draisma S, van Hemert AM, Schoevers RA, Kupka RW, Beekman ATF, et al. Depressive and anxiety disorders in concert–A synthesis of findings on comorbidity in the NESDA study. Journal of Affective Disorders. 2021;284:85–97.

23. Shao M, Lin X, Jiang D, Tian H, Xu Y, Wang L, et al. Depression and cardiovascular disease: Shared molecular mechanisms and clinical implications. Psychiatry Res. 2020;285:112802.

24. Tully PJ, Baumeister H, Martin S, Atlantis E, Jenkins A, Januszewski A, et al. Elucidating the Biological Mechanisms Linking Depressive Symptoms With Type 2 Diabetes in Men: The Longitudinal Effects of Inflammation, Microvascular Dysfunction, and Testosterone. Psychosom Med. 2016;78(2):221–32.

25. Baranova A, Liu D, Chandhoke V, Cao H, Zhang F. Unraveling the genetic links between depression and type 2 diabetes. Prog Neuropsychopharmacol Biol Psychiatry. 2025;137:111258.

26. Lim LF, Solmi M, Cortese S. Association between anxiety and hypertension in adults: A systematic review and meta-analysis. Neurosci Biobehav Rev. 2021;131:96–119.

27. Luo G, Li Y, Yao C, Li M, Li J, Zhang X. Prevalence of overweight and obesity in patients with major depressive disorder with anxiety: Mediating role of thyroid hormones and metabolic parameters. J Affect Disord. 2023;335:298–304.

28. Perry BI, Khandaker GM, Marwaha S, Thompson A, Zammit S, Singh SP, et al. Insulin resistance and obesity, and their association with depression in relatively young people: findings from a large UK birth cohort. Psychol Med. 2020;50(4):556–65.

29. Ninla-Aesong P, Puangsri P, Kietdumrongwong P, Jongkrijak H, Noipha K. Being overweight and obese increases suicide risk, the severity of depression, and the inflammatory response in adolescents with major depressive disorders. Front Immunol. 2023;14:1197775.

30. Mac Giollabhui N, Swistun D, Murray S, Moriarity DP, Kautz MM, Ellman LM, et al. Executive dysfunction in depression in adolescence: the role of inflammation and higher body mass. Psychol Med. 2020;50(4):683–91.

31. Khandaker GM, Pearson RM, Zammit S, Lewis G, Jones PB. Association of serum interleukin 6 and C-reactive protein in childhood with depression and psychosis in young adult life: a population-based longitudinal study. JAMA Psychiatry. 2014;71(10):1121–8.

32. Gomes AP, Gonçalves H, Dos Santos Vaz J, Kieling C, Rohde LA, Oliveira IO, et al. Do inflammation and adiposity mediate the association of diet quality with depression and anxiety in young adults? Clin Nutr. 2021;40(5):2800–8.

33. Hughes MF, Patterson CC, Appleton KM, Blankenberg S, Woodside JV, Donnelly M, et al. The Predictive Value of Depressive Symptoms for All-Cause Mortality: Findings From the PRIME Belfast Study Examining the Role of Inflammation and Cardiovascular Risk Markers. Psychosom Med. 2016;78(4):401–11.

34. Khandaker GM, Zuber V, Rees JMB, Carvalho L, Mason AM, Foley CN, et al. Shared mechanisms between coronary heart disease and depression: findings from a large UK general population-based cohort. Mol Psychiatry. 2020;25(7):1477–86.

35. Laake JP, Stahl D, Amiel SA, Petrak F, Sherwood RA, Pickup JC, et al. The association between depressive symptoms and systemic inflammation in people with type 2 diabetes: findings from the South London Diabetes Study. Diabetes Care. 2014;37(8):2186–92.

36. Wang Y, Liu M, Yang F, Chen H, Wang Y, Liu J. The associations of socioeconomic status, social activities, and loneliness with depressive symptoms in adults aged 50 years and older across 24 countries: findings from five prospective cohort studies. Lancet Healthy Longev. 2024;5(9):100618.

37. Eken HN, Dee EC, Powers AR, 3rd, Jordan A. Racial and ethnic differences in perception of provider cultural competence among patients with depression and anxiety symptoms: a retrospective, population-based, cross-sectional analysis. Lancet Psychiatry. 2021;8(11):957–68.

38. Vandenbroucke JP, von Elm E, Altman DG, Gøtzsche PC, Mulrow CD, Pocock SJ, et al. Strengthening the Reporting of Observational Studies in Epidemiology (STROBE): explanation and elaboration. Ann Intern Med. 2007;147(8):W163–94.

39. O’Bryant SE, Johnson LA, Barber RC, Braskie MN, Christian B, Hall JR, et al. The Health & Aging Brain among Latino Elders (HABLE) study methods and participant characteristics. Alzheimers Dement (Amst). 2021;13(1):e12202.

40. Yesavage JA, Brink TL, Rose TL, Lum O, Huang V, Adey M, et al. Development and validation of a geriatric depression screening scale: A preliminary report. Journal of Psychiatric Research. 1982;17(1):37–49.

41. Huang M, Su S, Goldberg J, Miller AH, Levantsevych OM, Shallenberger L, et al. Longitudinal association of inflammation with depressive symptoms: A 7-year cross-lagged twin difference study. Brain Behav Immun. 2019;75:200–7.

42. Lozupone M, Donghia R, Sardone R, Mollica A, Berardino G, Lampignano L, et al. Apolipoprotein E genotype, inflammatory biomarkers, and non-psychiatric multimorbidity contribute to the suicidal ideation phenotype in older age. The Salus in Apulia Study. J Affect Disord. 2022;319:202–12.

43. Elgellaie A, Thomas SJ, Kaelle J, Bartschi J, Larkin T. Pro-inflammatory cytokines IL-1α, IL-6 and TNF-α in major depressive disorder: Sex-specific associations with psychological symptoms. Eur J Neurosci. 2023;57(11):1913–28.

44. Amerikanou C, Valsamidou E, Kleftaki SA, Gioxari A, Koutoulogenis K, Aroutiounova M, et al. Peripheral inflammation is linked with emotion and mental health in people with obesity. A “head to toe” observational study. Front Endocrinol (Lausanne). 2023;14:1197648.

45. McLaughlin AP, Lambert E, Milton R, Mariani N, Kose M, Nikkheslat N, et al. Peripheral inflammation associated with depression and reduced weight loss: a longitudinal study of bariatric patients. Psychol Med. 2024;54(3):601–10.

46. Sominsky L, O’Hely M, Drummond K, Cao S, Collier F, Dhar P, et al. Pre-pregnancy obesity is associated with greater systemic inflammation and increased risk of antenatal depression. Brain Behav Immun. 2023;113:189–202.

47. McLachlan C, Shelton R, Li L. Obesity, inflammation, and depression in adolescents. Front Psychiatry. 2023;14:1221709.

48. Shelton RC, Falola M, Li L, Zajecka J, Fava M, Papakostas GI. The pro-inflammatory profile of depressed patients is (partly) related to obesity. J Psychiatr Res. 2015;70:91–7.

49. Ambrósio G, Kaufmann FN, Manosso L, Platt N, Ghisleni G, Rodrigues ALS, et al. Depression and peripheral inflammatory profile of patients with obesity. Psychoneuroendocrinology. 2018;91:132–41.

50. Wang K, Li F, Cui Y, Cui C, Cao Z, Xu K, et al. The Association between Depression and Type 1 Diabetes Mellitus: Inflammatory Cytokines as Ferrymen in between? Mediators Inflamm. 2019;2019:2987901.

51. Nguyen MM, Perlman G, Kim N, Wu CY, Daher V, Zhou A, et al. Depression in type 2 diabetes: A systematic review and meta-analysis of blood inflammatory markers. Psychoneuroendocrinology. 2021;134:105448.

52. Liu D, McIntyre RS, Li R, Yang M, Xue Y, Cao B. Genetic association between major depressive disorder and type 2 diabetes mellitus: Shared pathways and protein networks. Prog Neuropsychopharmacol Biol Psychiatry. 2021;111:110339.

53. Panagi L, Poole L, Steptoe A, Hackett RA. Inflammatory stress responses and future mental health outcomes in people with type 2 diabetes. Brain Behav Immun Health. 2022;23:100472.

54. Herder C, Schmitt A, Budden F, Reimer A, Kulzer B, Roden M, et al. Longitudinal associations between biomarkers of inflammation and changes in depressive symptoms in patients with type 1 and type 2 diabetes. Psychoneuroendocrinology. 2018;91:216–25.

55. Huang YQ, Wang Y, Hu K, Lin S, Lin XH. Hippocampal Glycerol-3-Phosphate Acyltransferases 4 and BDNF in the Progress of Obesity-Induced Depression. Front Endocrinol (Lausanne). 2021;12:667773.

56. Wu H, Lv W, Pan Q, Kalavagunta PK, Liu Q, Qin G, et al. Simvastatin therapy in adolescent mice attenuates HFD-induced depression-like behavior by reducing hippocampal neuroinflammation. J Affect Disord. 2019;243:83–95.

57. Meng L, Bai X, Zheng Y, Chen D, Zheng Y. Altered expression of norepinephrine transporter participate in hypertension and depression through regulated TNF-α and IL-6. Clin Exp Hypertens. 2020;42(2):181–9.

58. Alshogran OY, Khalil AA, Oweis AO, Altawalbeh SM, Alqudah MAY. Association of brain-derived neurotrophic factor and interleukin-6 serum levels with depressive and anxiety symptoms in hemodialysis patients. Gen Hosp Psychiatry. 2018;53:25–31.

59. van Sloten TT, Schram MT, Adriaanse MC, Dekker JM, Nijpels G, Teerlink T, et al. Endothelial dysfunction is associated with a greater depressive symptom score in a general elderly population: the Hoorn Study. Psychol Med. 2014;44(7):1403–16.

60. Gacar G, Gocmez SS, Halbutoğulları ZS, Kılıç KC, Kaya A, Yazir Y, et al. Resveratrol improves vascular endothelial dysfunction in the unpredictable chronic mild stress model of depression in rats by reducing inflammation. Behav Brain Res. 2023;438:114186.

61. The interleukin-6 receptor as a target for prevention of coronary heart disease: a mendelian randomisation analysis. The Lancet. 2012;379(9822):1214–24.

62. Interleukin-6 receptor pathways in coronary heart disease: a collaborative meta-analysis of 82 studies. The Lancet. 2012;379(9822):1205–13.

63. Euteneuer F, Neuert M, Salzmann S, Fischer S, Ehlert U, Rief W. Does psychological treatment of major depression reduce cardiac risk biomarkers? An exploratory randomized controlled trial. Psychol Med. 2023;53(8):3735–49.

64. Euteneuer F, Dannehl K, Del Rey A, Engler H, Schedlowski M, Rief W. Immunological effects of behavioral activation with exercise in major depression: an exploratory randomized controlled trial. Transl Psychiatry. 2017;7(5):e1132.

65. Hu MX, Penninx B, de Geus EJC, Lamers F, Kuan DC, Wright AGC, et al. Associations of immunometabolic risk factors with symptoms of depression and anxiety: The role of cardiac vagal activity. Brain Behav Immun. 2018;73:493–503.

66. Glaus J, Vandeleur CL, von Känel R, Lasserre AM, Strippoli MP, Gholam-Rezaee M, et al. Associations between mood, anxiety or substance use disorders and inflammatory markers after adjustment for multiple covariates in a population-based study. J Psychiatr Res. 2014;58:36–45.

67. Lamers F, Milaneschi Y, Vinkers CH, Schoevers RA, Giltay EJ, Penninx BWJH. Depression profilers and immuno-metabolic dysregulation: Longitudinal results from the NESDA study. Brain, Behavior, and Immunity. 2020;88:174–83.

68. Raison CL, Rutherford RE, Woolwine BJ, Shuo C, Schettler P, Drake DF, et al. A randomized controlled trial of the tumor necrosis factor antagonist infliximab for treatment-resistant depression: the role of baseline inflammatory biomarkers. JAMA Psychiatry. 2013;70(1):31–41.

69. McElvaney OJ, Curley GF, Rose-John S, McElvaney NG. Interleukin-6: obstacles to targeting a complex cytokine in critical illness. The Lancet Respiratory Medicine. 2021;9(6):643–54.

